# Digital PCR-based deep quantitative profiling delineates heterogeneity and evolution of uveal melanoma

**DOI:** 10.1101/2024.01.30.24301871

**Authors:** R.J. Nell, M. Versluis, N.V. Menger, R.M. Verdijk, W.G.M. Kroes, H.W. Kapiteijn, G.P.M. Luyten, M.J. Jager, P.A. van der Velden

## Abstract

Uveal melanoma is an aggressive intraocular tumour characterised by a limited number of genetic alterations. However, the evolution of this malignancy remains enigmatic. In this study, we performed a deep quantitative analysis of 80 primary uveal melanomas by novel digital PCR-based approaches. Mutations were quantified by targeted and drop-off mutation assays, copy number alterations were precisely measured by quantifying the allelic imbalance of heterozygous single-nucleotide polymorphisms. By comparing the absolute abundances of genetic alterations present in a bulk tumour, the heterogeneity and early evolution could be inferred. Tumour progression was further studied by analysing matched primary and metastatic lesions from five patients.

Gα_q_ signalling mutations were generically and always clonally present, suggesting to be acquired in the earliest stage of uveal melanoma development (‘primary driver’). Next, three main evolutionary subtypes could be identified based on having an *EIF1AX* mutation, *SF3B1* mutation or monosomy 3p. These alterations were usually mutually-exclusive and (near-) clonally abundant, suggesting to represent distinct secondary drivers. This contrasts with gains and amplifications of chromosome 8q, which were not restricted to one of the main subtypes and showed subclonality in 31% of the affected tumours. These tertiary alterations were not required for metastatic dissemination.

Using high-resolution analyses, we identified systematic differences in the evolutionary timing of genetic events in uveal melanoma. The observed intratumour heterogeneity suggests a more complex model of gradual tumour evolution and argues for a comprehensive genetic analysis in clinical practice, which may be facilitated by the sensitive digital PCR assays developed in this study.

## Introduction

Uveal melanoma (UM) is a rare, but often lethal tumour originating from malignantly transformed melanocytes in the choroid, ciliary body or iris in the eye [1, 2]. Unlike other types of melanomas, UMs present with a limited mutational burden and low number of structural variants [3, 4]. Nearly all tumours have a mutation in *GNAQ*, *GNA11*, *PLCB4* or *CYSLTR2*, which activates the Gα_q_ signalling pathway [5–8]. Most melanomas also harbour a so-called BSE mutation in *BAP1*, *SF3B1* or *EIF1AX* [3, 9–11]. The loss of chromosome 3p and the gain (i.e. one extra copy) or amplification (i.e. two or more extra copies) of chromosome 8q are the most prevalent copy number alterations (CNAs), and occur in typical combinations with the BSE mutations [12]. These distinct molecular subtypes are associated with specific gene expression profiles and are differentially correlated with the risk of metastatic spreading and patient survival [12–14].

Despite the large number of tumours studied, the evolution of UM remains incompletely understood [15–17]. Based on comprehensive bioinformatic analyses, UM has been proposed to arise after an early punctuated burst of the canonical genetic alterations, leading to relatively homogeneous tumours (i.e. most alterations are present in all malignant cells) [15, 17]. However, as most genomic data on UM have been generated by DNA sequencing of low to moderate depths, and the number of mutations is minimal in most tumours, these datasets may lack the power to reliably infer subclonal structures of heterogeneous tumours (i.e. alterations are present in varying proportions of the malignant cells) [18]. Recent work studying UMs in higher resolution (i.e. analyses at the level of single cells or of distinct progression stages) revealed a larger genomic complexity than appreciated previously [16, 19]. Though, these studies were carried out in relatively small cohorts or focussed specifically on the evolution of metastatic tumours, leaving the early evolution of primary UM relatively understudied. Moreover, as they were performed using advanced technical approaches, routine application of these techniques remains unavailable in research or clinical diagnostics.

As an alternative approach, a deep quantitative profiling of individual bulk DNA samples may provide insights into the heterogeneity and evolution of tumours, as we previously demonstrated in familial melanoma and *CYSLTR2* mutant UM [20, 21]. By determining the exact abundance of mutations and CNAs present in the bulk of a tumour, the inferred clonality of these alterations can be used to reconstruct the tumour’s genetic history: those alterations that are present in all malignant cells (‘clonal’) occurred early during tumour evolution, while events present in a subpopulation of the cells (‘subclonal’) occurred later during progression of the tumour.

In this study, we perform such in-depth characterisation of 80 primary UMs and five matched metastatic lesions using innovative digital PCR-based assays targeting the key genetic alterations of this malignancy. Hereby, we aim to delineate the heterogeneity of tumours and infer trends in the early and late evolution of UM.

## Materials and methods

### LUMC cohort sample collection and DNA/RNA isolation

Primary tumour specimens from 80 fresh-frozen UMs were collected from the biobank of the Department of Ophthalmology, Leiden University Medical Center (LUMC). All samples were obtained from patients primarily treated by enucleation between 2001 and 2011 in the LUMC. In addition, we collected five formalin-fixed and paraffin embedded (FFPE) biopsy specimens from tumour-matched metastatic lesions. This study was approved by the LUMC Biobank Committee and Medisch Ethische Toetsingscommissie under no. B14.003/DH/sh and B20.026. An overview of the clinical characteristics of all cases analysed in this study is presented in **Supplementary Table 1**.

DNA and RNA were isolated from 25x 20 µm sections using the QIAamp DNA Mini Kit and RNeasy Mini Kit respectively (Qiagen, Hilden, Germany). Total nucleic acid from FFPE tissue was isolated 4x 10 µm macrodissected sections using the Siemens Tissue Preparation System (Siemens Healthcare, Erlangen, Germany) [22]. For all procedures, the manufacturer’s instructions were followed.

### RNA sequencing analysis

We sequenced and analysed 100 ng of RNA per primary tumour sample as described previously [23]. In summary, samples were prepared using the NEBNext Ultra Directional RNA Library Prep Kit for Illumina (New England Biolabs, Ipswich, USA) and sequencing was performed using the Illumina HiSeq 4000 (Illumina, San Diego, USA) by GenomeScan (Leiden, the Netherlands). HiSeq control and image analysis, base calling and quality check were carried out using HCS (version 3.4.0), the Illumina data analysis pipeline RTA (version 2.7.7) and Bcl2fastq (version 2.17). Reads were aligned using STAR (version 2.5.3a) to human reference genome GRCh38. Gene counts were generated with htseq-count (version 0.6.0) and annotated using Ensembl (version 87). DESeq2 (version 1.30.0) was used for variance stabilising transformation of the count data. All aligned RNA sequencing files were screened for (hotspot) mutations in *GNAQ* (p.Q209, p.R183 and p.G48), *GNA11* (p.Q209, p.R183), *CYSLTR2* (p.L129), *PLCB4* (p.D630), *EIF1AX* (entire gene), *SF3B1* (exon 14) and *BAP1* (entire gene) using freebayes (version 1.3.6), and via manual inspection using the Integrative Genomics Viewer (version 2.8.10). The aligned RNA sequencing files were further analysed for single-nucleotide polymorphisms (SNPs) using VarScan (version 2.3), which were annotated using snpEff/snpSift (version 5.1) according to dbSNP (version 151). Only heterozygously and highly expressed common variants (flagged as ‘common’, total number of reads > 40, number of reference reads > 2, number of alternate reads > 2) were included in the downstream analysis to identify CNA-derived allelic imbalances.

### Digital PCR experiments and integration of results

Digital PCR experiments were carried out using the QX200 Droplet Digital PCR System (Bio-Rad Laboratories, Hercules, USA) following the protocols and general guidelines described and discussed earlier [21, 24–27]. In general, 20 ng DNA was analysed in a 22 uL experiment, using 11 uL ddPCR™ Supermix for Probes (No dUTP, Bio-Rad) and primers and probes in final concentrations of 900 and 250 nM for duplex experiments, or optimised concentrations for multiplex experiments. The context sequence, PCR annealing temperature and supplier information for all assays used is provided in **Supplementary Table 2**.

PCR mixtures were partitioned into 20.000 droplets using the AutoDG System (Bio-Rad). The PCR was performed in a T100 Thermal Cycler (Bio-Rad) with the following protocol: 10 min at 95 °C; 30 s at 94 °C and 1 min at 55 °C, 58 °C or 60 °C (depending on the assay, see **Supplementary Table 2**) for 40 cycles; 10 min at 98 °C; cooling at 12 °C for up to 48 h, until droplet reading. The ramp rate was set to 2 °C/s for all steps. A QX200 Droplet Reader (Bio-Rad) was used to perform reading of the droplets.

Raw digital PCR results were acquired using QuantaSoft (version 1.7.4, Bio-Rad) and imported and analysed in the online digital PCR management and analysis application Roodcom WebAnalysis (version 2.0, available via https://webanalysis.roodcom.nl).

The Gα_q_ signalling mutations (in *GNAQ*, *GNA11*, *CYSLTR2* and *PLCB4*) and BSE mutations (in *BAP1*, *SF3B1* and *EIF1AX*) were analysed in duplex digital PCR experiments and the measured concentrations (listed between square brackets) of the mutant and wild-type alleles were used to calculate the mutant allele fractions:

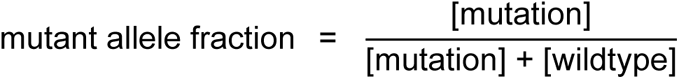

These values were then used to estimate the mutant cell fractions under heterozygous conditions:

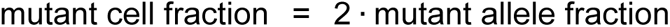

In line with our recent study [27], two approaches were employed to measure CNAs. Firstly, following the classic approach, copy number values were calculated based on the measured concentrations of a DNA target and one stable diploid reference gene:

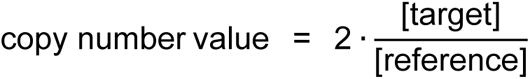

The targets included genes on chromosome 3p (*PPARG*) and 8q (*PTK2*). Genes on chromosome 5p (*TERT*), 7p (*VOPP1*), 7q (*BRAF*) or 14q (*TTC5*) were evaluated as stable references. Moreover, we used this approach to evaluate the copy number values of the mutant and wild-type alleles in the case of an allelic imbalance affecting the Gα_q_ signalling and BSE genes. As described earlier [21], these additional measurements allowed to calculate the mutant cell fractions under non-heterozygous conditions:

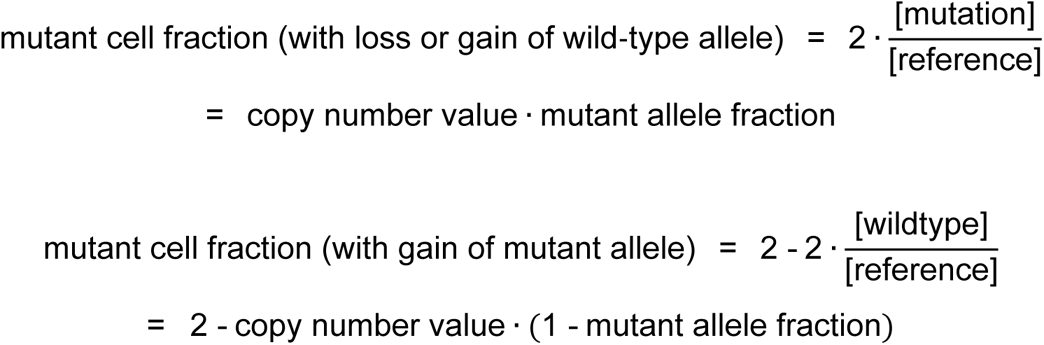

Secondly, following the SNP-based approach, copy number values were measured using the concentrations of haploid genomic loci var_1_ and var_2_, representing the two variants of a heterozygous SNP:

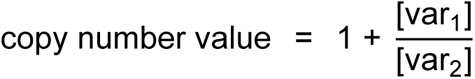

In this formula, the SNP variant in the denominator (var_2_) is the one assumed to be unaltered and copy number stable (i.e. haploid). When measuring (potential) chromosomal losses, the variant with the highest concentration was chosen as stable variant. Vice versa, the variant with the lowest concentration was assumed to be stable when analysing chromosomal gains and amplifications. Analysed SNPs in this study included rs6617, rs6976, rs9586, rs1989839, rs1062633 and rs2236947 on chromosome 3p, and rs7018178 and rs7843014 on chromosome 8q.

The clonality of the genetic alterations was primarily assessed by comparing the mutant cell fractions in individual tumours. In most samples, the Gα_q_ signalling mutation represented the largest and thus clonal fraction of cancer cells and was considered an accurate estimate of the total fraction malignant cells (i.e. tumour purity). The clonality of CNAs was determined by adjusting the measured copy number values for this tumour purity (assuming that admixed non-malignant cells always have a copy number value of 2). Non-integral outcomes were considered subclonal. For simple CNAs (between −1 and +1), the quantified loss or gain can be mathematically attributed to a specific proportion of the cells in a tumour [21]:

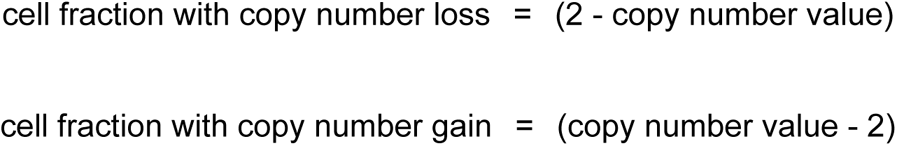

For all digital PCR experiments, 99%-confidence intervals were calculated following Fieller’s theorem and taken into account in all calculations [28]. *P* values < 0.01 were considered statistically significant.

### Validation of findings with karyotyping and BAP1 immunohistochemistry

In a selection of cases, the heterogeneity of CNAs and presence and pathogenicity of *BAP1* alterations were confirmed by karyotyping and an immunohistochemical staining, both performed as routine diagnostic analyses in two ISO accredited laboratories (Departments of Pathology, LUMC) [25, 29, 30].

### Bioinformatic analyses and data availability

The bioinformatic analyses were carried out using R (version 4.0.3) and RStudio (version 1.4.1103). Custom scripts and unprocessed results are available via https://github.com/rjnell/um-heterogeneity. The molecular interpretation of all cases analysed in this study is presented in **Supplementary Table 1**.

## Results

### Sample collection and RNA sequencing analysis

Primary UM specimens from 80 patients were studied for mutations and CNAs at the DNA level using digital PCR. As this is a largely targeted technique, RNA sequencing was performed to screen for the presence of genetic alterations, an approach we recently validated for UM [23]. In addition, RNA sequencing was used to divide our cohort into class I (n=29) or class II (n=51) tumours, based on their bulk gene expression profile (GEP). An overview of the molecular and clinical characteristics of all cases is presented in **Figure 1 and Supplementary Table 1**.

**Figure 1.**
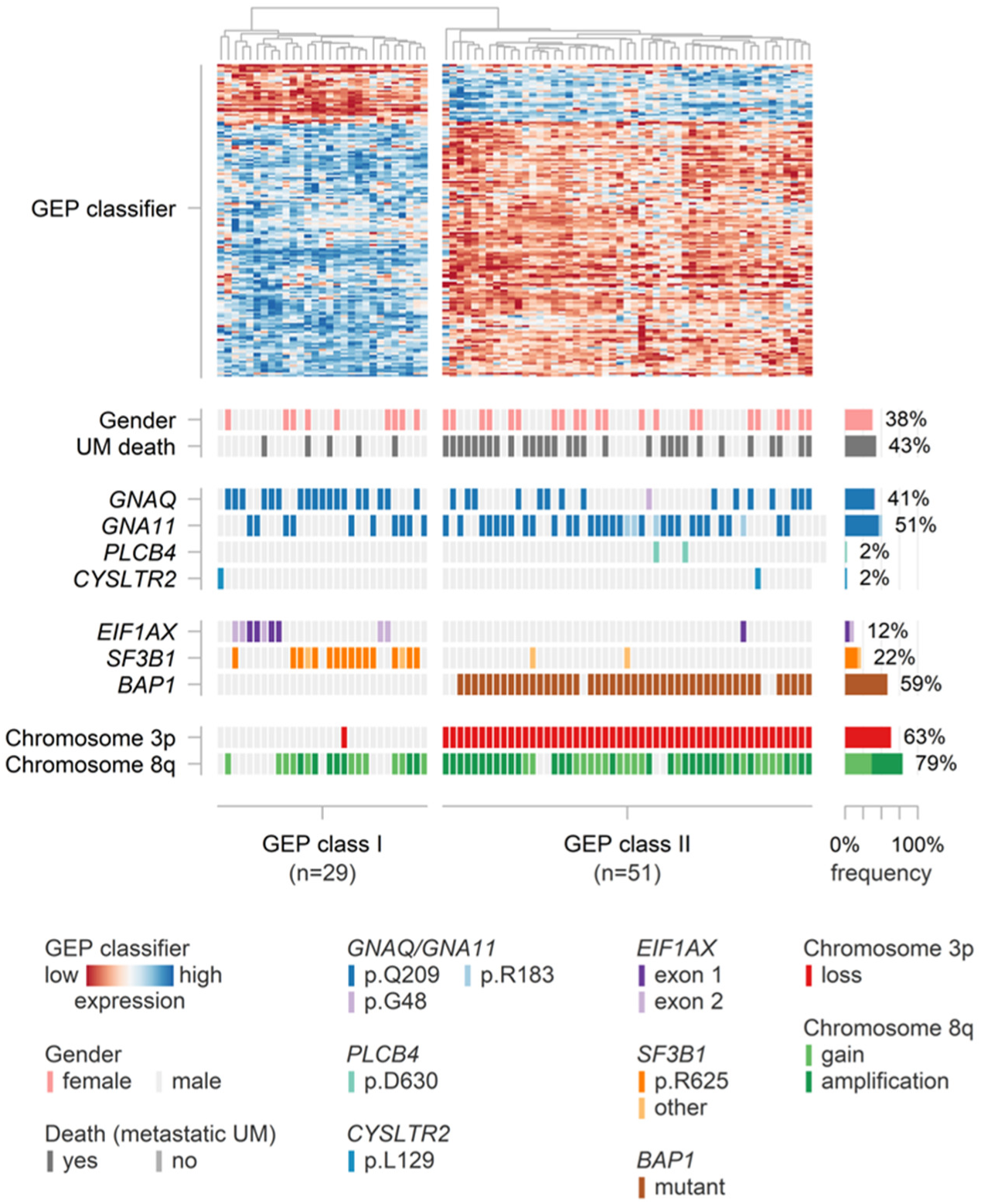
Overview of the GEP classification, clinical characteristics and genetic alterations in our LUMC cohort of primary UMs.

### Gα_q_ and BSE mutation analysis

Gα_q_ signalling mutations were detected in 79/80 (99%) tumours of the LUMC cohort and analysed at the DNA level via accustomed targeted digital PCR assays (**Figure 2A and Supplementary Table 1**) Although the mutations recurrently affected the established hotspot positions (p.Q209 and p.R183 in *GNAQ*/*GNA11*, p.D630 in *PLCB4* and p.L129 in *CYSLTR2*), considerable variation was present at the nucleotide level (**Figure 2B**). The *GNAQ* p.G48L mutation, which seems to represent a rare alternative hotspot [12, 16, 31, 32], was identified in one tumour.

**Figure 2.**
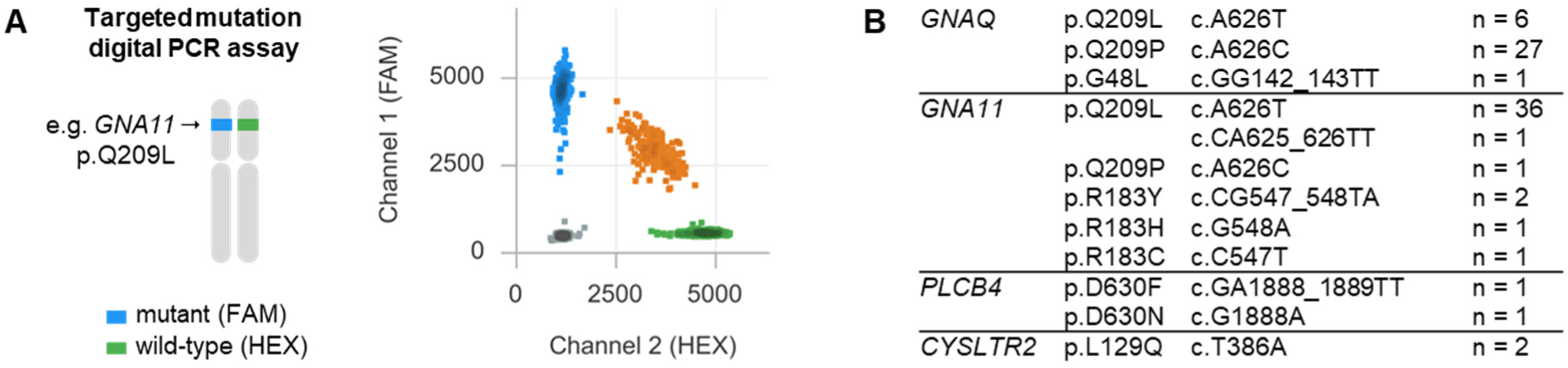
(**A**) Analysis of hotspot Gα_q_ signalling mutations using duplex digital PCR with FAM- and HEX-labelled probes targeting the mutant and wild-type sequences. (**B**) Prevalence and exact nucleotide variation of the Gα_q_ signalling mutations in the LUMC cohort.

*EIF1AX* mutations are known to affect the two first exons of the gene [11], and were detected using RNA sequencing in 11/80 (14%) cases (**Figure 3A and Supplementary Table 1**). To verify these mutations at the DNA level, we developed two drop-off digital PCR assays. These assays were designed to target a polymorphic wild-type sequence using two fluorophore-labelled probes within one PCR amplicon [33]. When wild-type alleles are present, a double-positive signal is measured, while when a mutation is present, one of the two probes ‘drops off’ and single-positive droplets appear (**Figure 3B**). This way, we were able to confirm and quantify the presence of all 11 mutations using only two generic assays.

**Figure 3.**
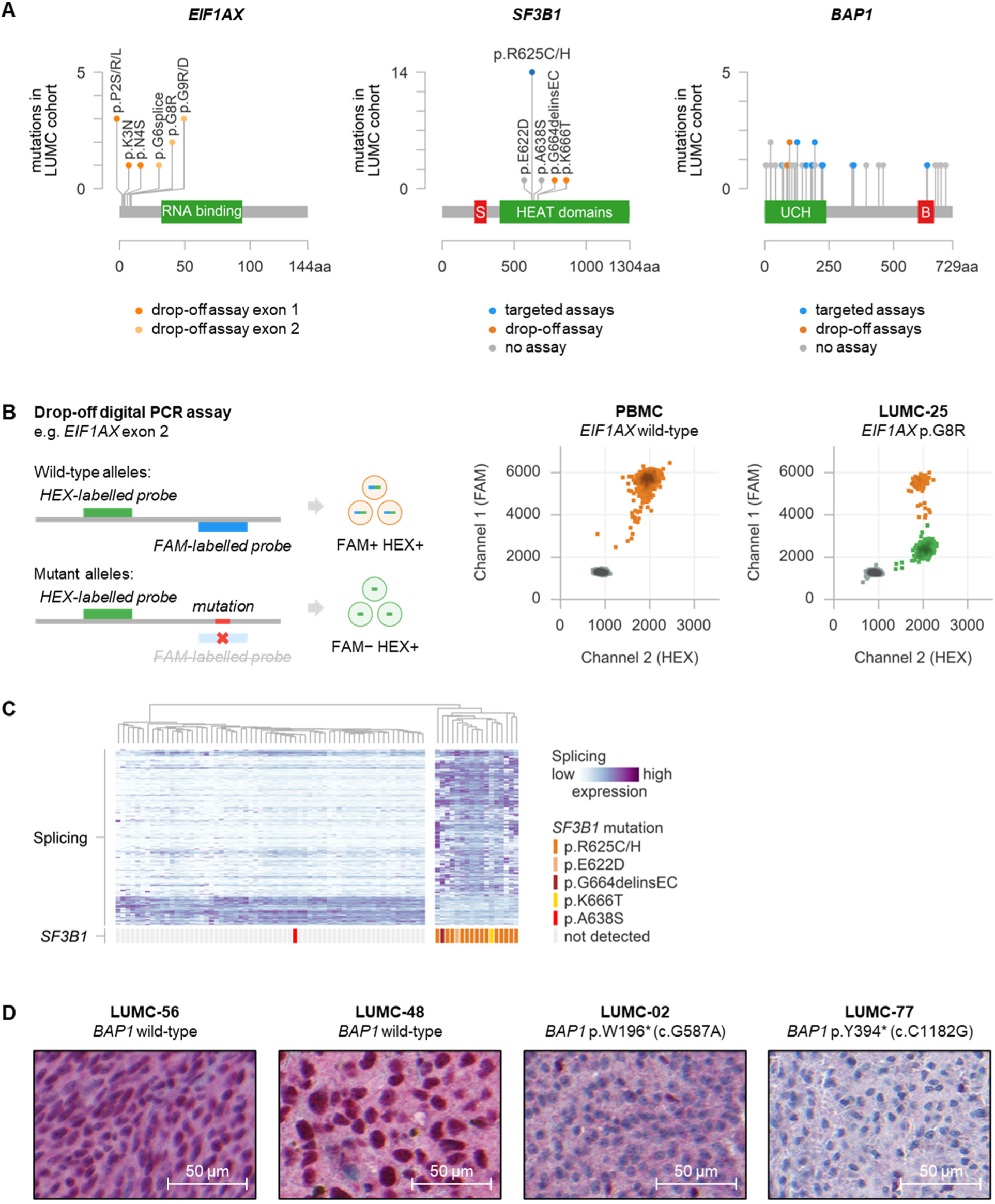
(**A**) Lollipop plots demonstrating the distribution and prevalence of mutations in *EIF1AX*, *SF3B1* and *BAP1* in the LUMC cohort. The colors refer the digital PCR assays available to analyse these mutations. Red and green boxes refer to selected gene domains. (**B**) Illustration of the principle behind the digital PCR drop-off assays with two 2D plots as examples of the *EIF1AX* exon 2 drop-off analysis. (**C**) Analysis of a signature of alternative splicing to identify *SF3B1* mutant tumours. (**D**) Representative immunohistochemical BAP1 nuclear stainings demonstrating preserved staining in *BAP1* wild-type tumours and lost staining in tumours with a *BAP1* mutation.

*SF3B1* (Splicing Factor 3B subunit 1A) encodes for a component of the splicing machinery and mutations in this gene result in aberrant splicing [3, 34]. Focussing on the exon 14 hotspot region, RNA sequencing identified *SF3B1* mutations in 18/80 (23%) melanomas, with 14 cases affecting the p.R625 position (**Figure 3A and Supplementary Table 1**).

*SF3B1*-mutation-associated deregulated splicing was observed in 17/18 mutant cases, while this was not detected in any of the *SF3B1*-wild-type tumours (**Figure 3C**). The only discordant case (LUMC-79) harboured a novel *SF3B1* p.A638S mutation of unknown pathogenic significance. The *SF3B1* mutant allele fractions were quantified at the DNA level in 16 cases using digital PCR assays targeting the p.R625C or p.R625H mutation and a dedicated drop-off assay.

Inactivating *BAP1* mutations (or deletions) can be found throughout the complete *BAP1* gene, challenging the identification of all of these alterations using conventional sequencing [15, 35]. RNA sequencing of our cohort helped us to identify 43/80 (55%) tumours with various types of probably damaging *BAP1* alterations (**Figure 3A and Supplementary Table 1**). In five tumours, the sequencing reads insufficiently covered the entire gene, and no conclusions could be drawn on the presence of any mutation. For a selection of cases, an immunohistochemical BAP1 protein staining confirmed the presence or absence of *BAP1* inactivating alterations (**Figure 3D and Supplementary Table 1**). To analyse the exact abundance and clonality of a variety of *BAP1* mutations, we developed one drop-off and eight targeted digital PCR assays by which 15 tumours from our cohort could be characterised.

### Allele-specific copy number analysis of chromosome 3p and 8q alterations

Extending our recent proof-of-concept study [27], chromosome 3p and 8q CNAs were analysed using two complementary digital PCR-based approaches. For the classic approach, targets on chromosome 3p and 8q were quantified against a stable reference gene using multiplex digital PCR. As a reference-independent alternative, CNA-derived allelic imbalances were measured by quantifying the two variants of heterozygous common SNPs on chromosome 3p and 8q in duplex digital PCR experiments.

Generally, both approaches resulted in highly comparable copy number values (Spearman’s rho = 0.95, *p* < 0.001, **Figure 4A**). By integrating the values determined with the classic and SNP-based approaches, copy number stability of one allele (i.e. one SNP variant) could be assured in most tumours (**Figure 4B**). This confirms our earlier observation that CNAs affecting chromosomes 3p and 8q usually involve only one of both alleles [27]. In our current cohort, we did not identify cases with isodisomy 3p, which would have been visible as a large allelic imbalance in combination with an absent or limited absolute loss of chromosome 3p. Moreover, in 49/55 (89%) of the informative tumours, the chromosome 8q copy number value could be correctly estimated based on the allelic (im)balance, which required genomic stability of at least one of both alleles (i.e. one SNP variant). This indicates that 8q copy number increases, including the larger amplifications, are predominantly derived from the (consecutive) gain of the maternal or paternal allele solely (**Figure 4B**). In six tumours we observed that the amplification originated from gaining both alleles. Since the allele-specific copy numbers were significantly different in all these cases (**Supplementary Figure 1A**), these amplifications are likely the result of separate genetic events. As representatively illustrated by case LUMC-75, for which we integrated our copy number data with the allelic imbalances observed in the RNA sequencing data [23], a multi-chromosomal or genome-wide ploidy change could be excluded, indicating that the amplification was acquired via an 8q-specific mechanism (**Supplementary Figure 1B**). In 31 tumours, chromosome 3p and/or 8q copy numbers could be determined via multiple heterozygous SNPs, which further validated the technical reproducibility of our assays and SNP-based approach (**Supplementary Figure 1C**). As this approach – in line with our recent study [27] – also led to more precise copy number measurements (**Supplementary Figure 1D**), we focussed on the SNP-based results in downstream evaluation of all copy numbers.

**Figure 4.**
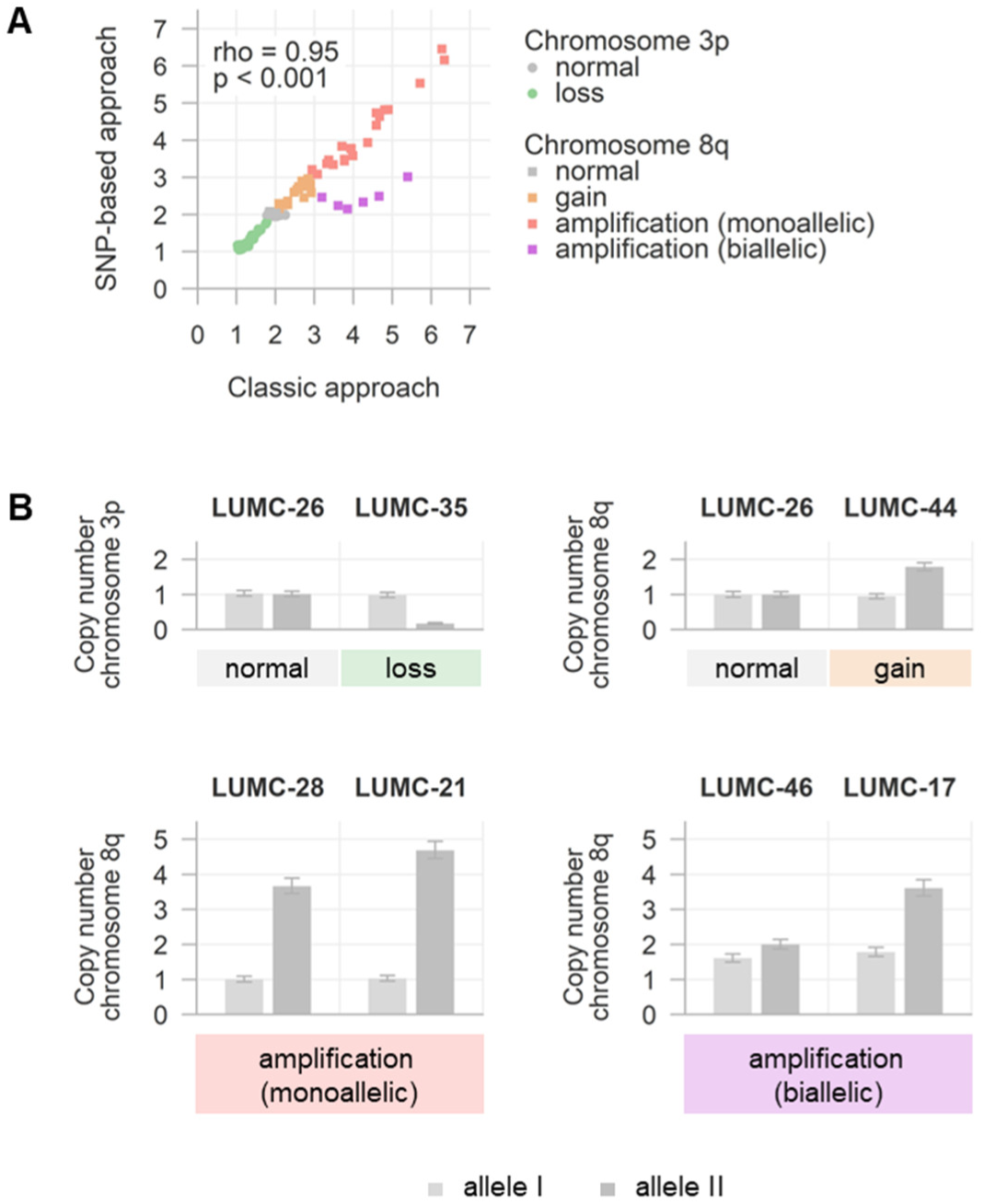
(**A**) Comparison of chromosome 3p and 8q copy number values determined with the classic and SNP-based approaches. (**B**) Examples of integrated copy number values determined with both approaches, revealing the allele-specificity behind the various alterations.

### Clonality analysis integrating mutations and CNAs

By comparing the absolute abundances of the somatic genetic alterations present in a bulk tumour, the clonality and heterogeneity of individual cases can be inferred [36–39]. For this goal, we integrated the quantified mutant allele fractions with the copy number states of the respective genomic loci (indicating zygosity of the mutations, see **Methods**) to determine the Gα_q_, *EIF1AX*, *SF3B1* and *BAP1* mutant cell fractions (**Figure 5A**). Through adjusting all copy number values for tumour purity (i.e. the fraction malignant cancer cells in a tumour sample), the clonality of the various CNAs could also be assessed (**Figure 5A**). This analysis was based on the assumption that CNAs – when clonally present in all malignant cells – should give an integral copy number: −1 for a loss, +1 for a gain and +2, +3, +4 et cetera for an amplification. When the copy number is significantly different from these integral values, the alteration can only originate from a mixture of subpopulations, as exemplified by comparing our bulk measurements to single-cell karyotyping (**Figure 5B**, complete karyograms are presented in **Supplementary Figure 2**). For simple CNAs (between −1 and +1), the quantified loss or gain can be mathematically attributed to a certain proportion of the tumour cells.

**Figure 5.**
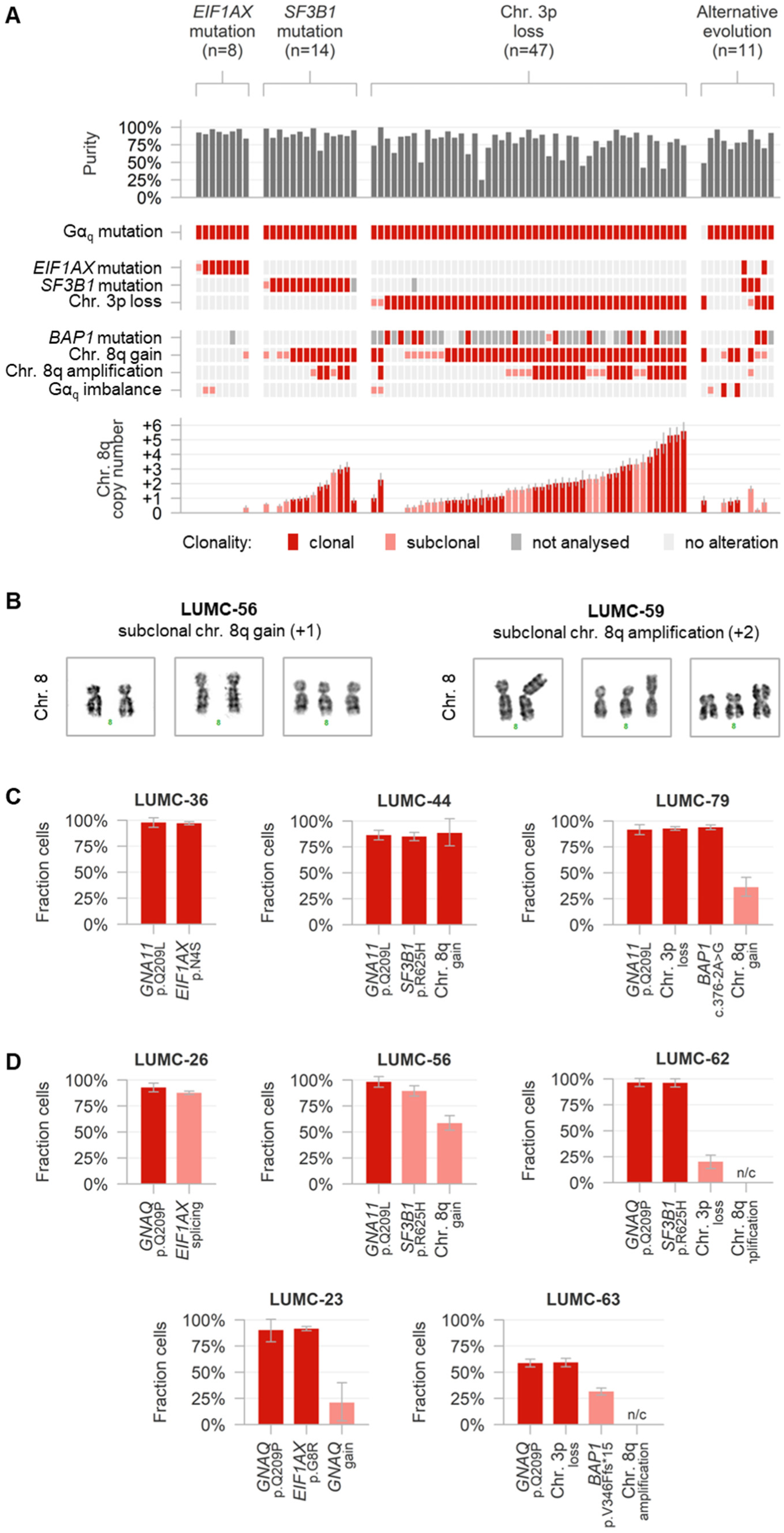
(**A**) Overview of presence, clonality and mutual-exclusivity of the various genetic alterations in the LUMC cohort. Additionally, the measured tumour purities and purity-corrected chromosome 8q copy number values are presented (**B**) Evidence of chromosome 8q heterogeneity by single-cell karyotyping. (**C**) Examples of tumours with multiple clonal genetic alterations. (**D**) Examples of tumours with a clonal Gα_q_ signalling mutation and additional (sub)clonal alterations. N/c: not calculable.

A total of 79/80 primary UMs of our cohort carried a Gα_q_ signalling mutation, which was always clonally expanded (**Figure 5C-D)**. Although the Gα_q_ mutations were typically heterozygous, in 8/79 (10%) tumours an additional CNA altered the absolute or relative dosage of the mutant allele. This involved loss of wild-type *GNAQ* (n=3), loss of wild-type *GNA11* (n=1), gain of mutant *GNAQ* (n=1) and gain of wild-type *GNAQ* (n=1), next to the cases with loss of wild-type *CYSLTR2* (n=1) and gain of mutant *CYSLTR2* (n=1) described previously [21]. These additional alterations were subclonal in 6/8 tumours (**Figure 5E**).

Mutations in *EIF1AX* were found in 10 melanomas and were clonally present in the entire population of malignant cells in nearly all cases (**Figure 5C**). Only one tumour showed a small but significantly detectable fraction of Gα_q_ mutant malignant cells that were *EIF1AX* wild-type (*p* < 0.01, **Figure 5D**). As *EIF1AX* is located on chromosome X, differences were observed in the abundances of mutant and wild-type alleles between female (n=1, heterozygous mutation) and male patients (n=10, hemizygous mutation), but in none of the tumours additional CNAs involving this gene were identified.

*SF3B1* mutations were found in 18 tumours, of which 16 could be further analysed and also tended to be clonally present (**Figure 5C**). In only one case a small fraction of *SF3B1* wild-type malignant cells (*p* < 0.01, **Figure 5D**) was measured. No CNAs affecting mutant or wild-type *SF3B1* were identified.

Loss of chromosome 3p was observed in 52/80 (65%) of the tumours. This monosomy was clonally expanded in most tumours (**Figure 5C**). Two GEP class II tumours presented with a small fraction of disomic malignant cells, of which one was *CYSLTR2* mutant and described in our earlier study [21]. Of the GEP class I tumours, only one showed monosomy, which turned out to be subclonally present in a larger fraction of the malignant cells (*p* < 0.01, **Figure 5D**).

Pathogenic *BAP1* alterations were detected in 46 tumours (all presenting with monosomy 3p) and targeted analysis in a selection of 15 tumours confirmed a clonal inactivation of *BAP1* in 14 cases: both the mutation and the loss of the other chromosome 3p were present in the complete Gα_q_ mutant malignant cell fraction (**Figure 5C**). In one tumour, we observed that the *BAP1* mutation was only present in a subclone (*p* < 0.01), while the monosomy 3p was clonally present in all malignant cells (**Figure 5D**).

Only 15/80 (19%) tumours of our cohort had a normal copy number of chromosome 8q. In those tumours with a detected gain or amplification, large variation was observed in the absolute numbers of additional copies (**Figure 5A**). In total, subclonal gains were found in 13 tumours, clonal gains in 17 tumours, subclonal amplifications in 12 tumours and clonal amplifications in 23 tumours.

### Identification of three main UM subtypes based on clonality and mutual-exclusivity

Taken together, a single clonal Gα_q_ signalling mutation was found in nearly all studied tumours, making it the most generic genetic event in our cohort. Though, most UMs acquired additional alterations (**Figure 5A**). Based on trends in their clonality and mutual-exclusivity, three main subgroups of tumours could be defined by (1) a disomy 3p with an *EIF1AX* mutation (8/80, 10% of our cohort), (2) a disomy 3p with an *SF3B1* mutation (14/80, 18%), or (3) a monosomy 3p (47/80, 59%). As these genetic alterations were all (near-)clonally abundant, it is likely that they occurred early during UM development.

In contrast, chromosome 8q CNAs were not restricted to one of these three subtypes nor obligately present in all tumours of one subtype. Moreover, these copy number increases frequently showed subclonality (**Figure 5A**). This indicates that – in contrast to the clonal alterations – 8q copy number increases were only present in subpopulations of the malignant cells and were acquired at a later evolutionary time point.

When the loss of chromosome 3p is accompanied by a mutation in *BAP1*, a complete inactivation of this tumour suppressor gene can be accomplished. Similar to the heterogeneity of chromosome 8q CNAs – *BAP1* alterations were not detected in all monosomic tumours of our cohort, BAP1 protein expression was not always lost and the *BAP1* mutation was not always clonally expanded (**Figure 3D and 5A**).

The remaining cases (11/80, 14%) were collectively grouped for their ‘alternative’ profile of genetic alterations with none (n=1) or two clonal Gα_q_ mutations (n=1), a disomy 3p without any detectable BSE mutation (n=5) or with both a clonal *EIF1AX* and *SF3B1* mutation (n=1), a clonal monosomy 3p with both a clonal *BAP1* and *EIF1AX* mutation (n=1) or a clonal *BAP1* and *SF3B1* mutation (n=1), or a clonal *SF3B1* mutation with a subclonal monosomy 3p (n=1). In these tumours chromosome 8q gains and amplifications were also heterogeneously present.

Of note, the inferred tumour purity differed substantially between individual tumours (median 86%, range 25-100%), with the lowest fractions of malignant cells observed in UMs with loss of chromosome 3p (**Figure 5A and Supplementary Table 1**).

From a clinical perspective, univariate analyses showed that a larger basal diameter, a lack of *SF3B1* mutation, a loss of chromosome 3p and a higher 8q copy number were related to a worse melanoma-related survival (**Table 1**). Age, gender, tumour prominence and having an *EIF1AX* mutation did not reach statistical significance. In a multivariate analysis, only the largest basal diameter of the tumour and the loss of chromosome 3p remained significantly associated with a poor melanoma-related survival.

**Table 1.** Uni- and multivariate Cox proportional hazards analysis for factors associated with melanoma-related survival. HR = hazard ratio, vs. = versus, bold underlined *p* values are considered statistically significant (*p* < 0.05).

| Variable | Univariate |  | Multivariate |  |
| --- | --- | --- | --- | --- |
|  | HR | *p* value | HR | *p* value |
| Age | 1.001 | 0.941 | 0.988 | 0.397 |
| Gender (male versus female) | 0.855 | 0.652 | 1.160 | 0.680 |
| Tumour prominence (mm) | 1.019 | 0.744 | 0.979 | 0.742 |
| Tumour largest basal diameter (mm) | 1.143 | **<u>0.022</u>** | 1.149 | **<u>0.042</u>** |
| *EIF1AX* mutation (versus wildtype) | 0.185 | 0.096 | 0.474 | 0.526 |
| *SF3B1* mutation (versus wildtype) | 0.324 | **<u>0.035</u>** | 0.670 | 0.607 |
| Chromosome 3p loss (versus disomy) | 5.667 | **<u><0.001</u>** | 4.325 | **<u>0.040</u>** |
| Chromosome 8q (purity-corrected copy number) | 1.397 | **<u>0.002</u>** | 0.968 | 0.840 |

### Analysis of five metastases reveals parallel 8q evolution

To investigate the clonality and heterogeneity of the primary UMs in relation to metastatic progression, we extended our study with the genetic analysis of metastatic lesions from five patients of our cohort.

Several genetic alterations were shared between the primary tumours and matched metastases (**Figure 6A**). This included Gα_q_ signalling mutations in all patients, *SF3B1* mutations in LUMC-42 and -22, chromosome 3p losses in LUMC-29, -79 and -73, and *BAP1* mutations in LUMC-79 and -73. All these alterations were clonally abundant in both primary tumour and metastasis, suggesting to reside on the trunk of the evolutionary trees (**Figure 6B**).

**Figure 6.**
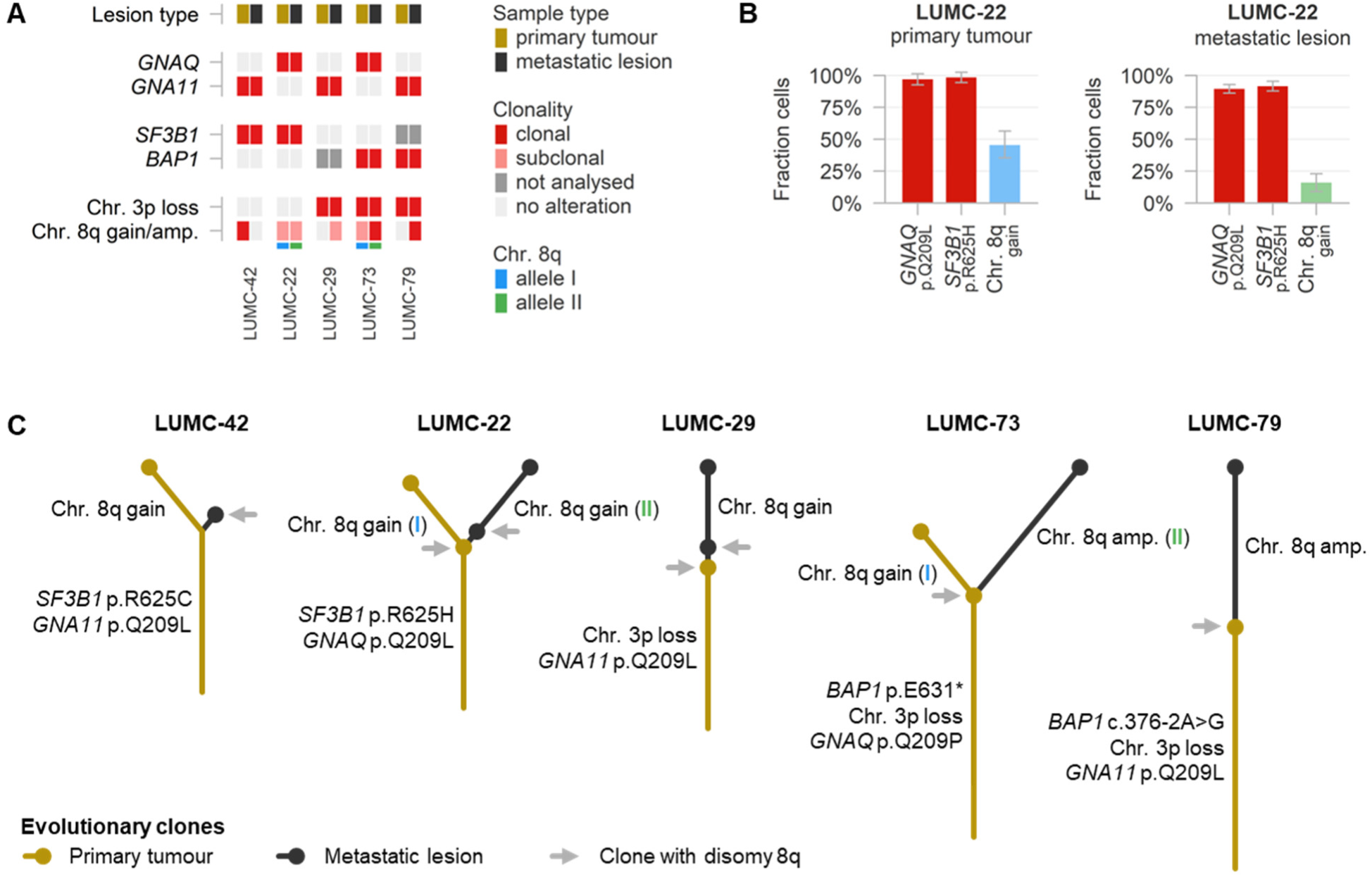
(**A**) Overview of genetic alterations and their clonality in five pairs of primary tumours and metastatic lesions. Additionally, the allele-specificity of the chromosome 8q CNAs is presented. (**B**) Extended from **A**. Detailed example of the clonality analysis in the samples from case LUMC-22. Note that the chromosome 8q gain is subclonally present in both samples, but involves a different allele. (**C**) Inferred evolutionary trees of the uveal melanomas from the five patients.

Regarding chromosome 8q, however, large variation was observed between the primary and metastatic lesions (**Figure 6A**). In LUMC-29 and -73 no chromosome 8q copy number increases were detected in the primary tumours, but the metastases presented with a subclonal gain or clonal amplification of this chromosome. In LUMC-42 the primary tumour had a clonal 8q gain, but the metastasis presented with a disomy 8q. In LUMC-22 and -79 a subclonal 8q gain was identified in the primary tumour, but the metastatic lesions harboured a subclonal gain and clonal amplification of the other allele of chromosome 8q (**Figure 6A and B**).

In aggregate, 8q copy number increases encompassed independent evolutionary events that could be placed on separate branches of the evolutionary trees (**Figure 6C**). In all patients, the metastatic clone was most likely derived from primary tumour cells carrying a panel of genetic alterations (Gα_q_ signalling mutation, *SF3B1* mutation, chromosome 3p loss and/or *BAP1* mutation), but a disomy 8q. Additional 8q copies were acquired later, during progression of primary tumour or metastasis (**Figure 6C**).

## Discussion

Cancer develops through the accumulation of somatic genetic alterations over time [40, 41]. The evolution of human malignancies, including various forms of melanoma, has been elucidated by comparing the genetic make-up of distinct progression stages of individual tumours [42–45]. In UM, however, tumour material is only scarcely available and histologically distinct precursor lesions next to a melanoma are rarely observed [1]. Consequently, the (early) evolution of this malignancy remains incompletely understood [15–17]. As an alternative approach, the evolutionary history of an individual tumour can be inferred by a deep quantitative assessment of the genetic alterations present in the bulk of a single tumour biopsy [36–39]. To better understand the development of UM, we performed such in-depth analysis by focussing on the most common mutations and CNAs in this malignancy. For this goal, we successfully screened RNA sequencing data from 80 primary UMs for their genetic alterations, and effectively applied novel, dedicated digital PCR assays to measure the clonality of these alterations at the DNA level.

In 99% of the tumours in our cohort, we identified an activating Gα_q_ signalling mutation in *GNAQ*, *GNA11*, *CYSLTR2* or *PLCB4*, confirming their ubiquitous presence in UM (**Figure 1**). We found (almost completely) mutually-exclusive mutations at all established hotspots, including the rare *GNAQ* p.G48 position (**Figure 2B**). As the Gα_q_ signalling mutations were always clonally present, they were likely acquired in one of the earliest stages of tumour development. The common presence of the same mutations in benign nevi and melanocytosis in the eye – both supposed precursors for UM – supports this hypothesis [21, 46, 47]. Consequently, a role in tumour initiation (‘primary driver’) rather than in malignant progression is postulated. Several studies, nevertheless, question the relation between mutations and activity of the Gα_q_ signalling genes and progression of UM [12, 48]. Additional genetic alterations have been described that elevated the allelic balance at the mutant locus towards the *GNAQ* or *CYSLTR2* mutations [16, 21]. We identified such alterations in 9% of the tumours in our cohort, and also one involving *GNA11*. Their typical presence in only a tumour subclone confirms that these alterations were acquired in a later phase, during progression of the primary tumour.

In addition to the early Gα_q_ signalling mutation, most UMs acquired additional genetic changes. These occurred in typical combinations and followed a recurring order of events, as summarised in **Figure 7A**. Three main evolutionary UM subtypes could be identified based on having an *EIF1AX* mutation, *SF3B1* mutation or monosomy 3p. These three alterations were usually mutually-exclusive and clonally abundant, suggesting to represent distinct ‘secondary drivers’. This finding matches with the paradigm that tumours with an *EIF1AX* mutation, *SF3B1* mutation or monosomy 3p represent distinct molecular entities and is in line with earlier reports of the *EIF1AX* and *SF3B1* mutations typically being clonally abundant [12, 15–17, 19]. However, in contrast to some previous studies [49, 50], we only rarely observed heterogeneity of monosomy 3p within the malignant cell population. We hypothesise that the low tumour purity of this latter subtype, sometimes even lower than 50% (**Figure 5A**), might explain why disomic and monosomic cells have frequently been found together.

**Figure 7.**
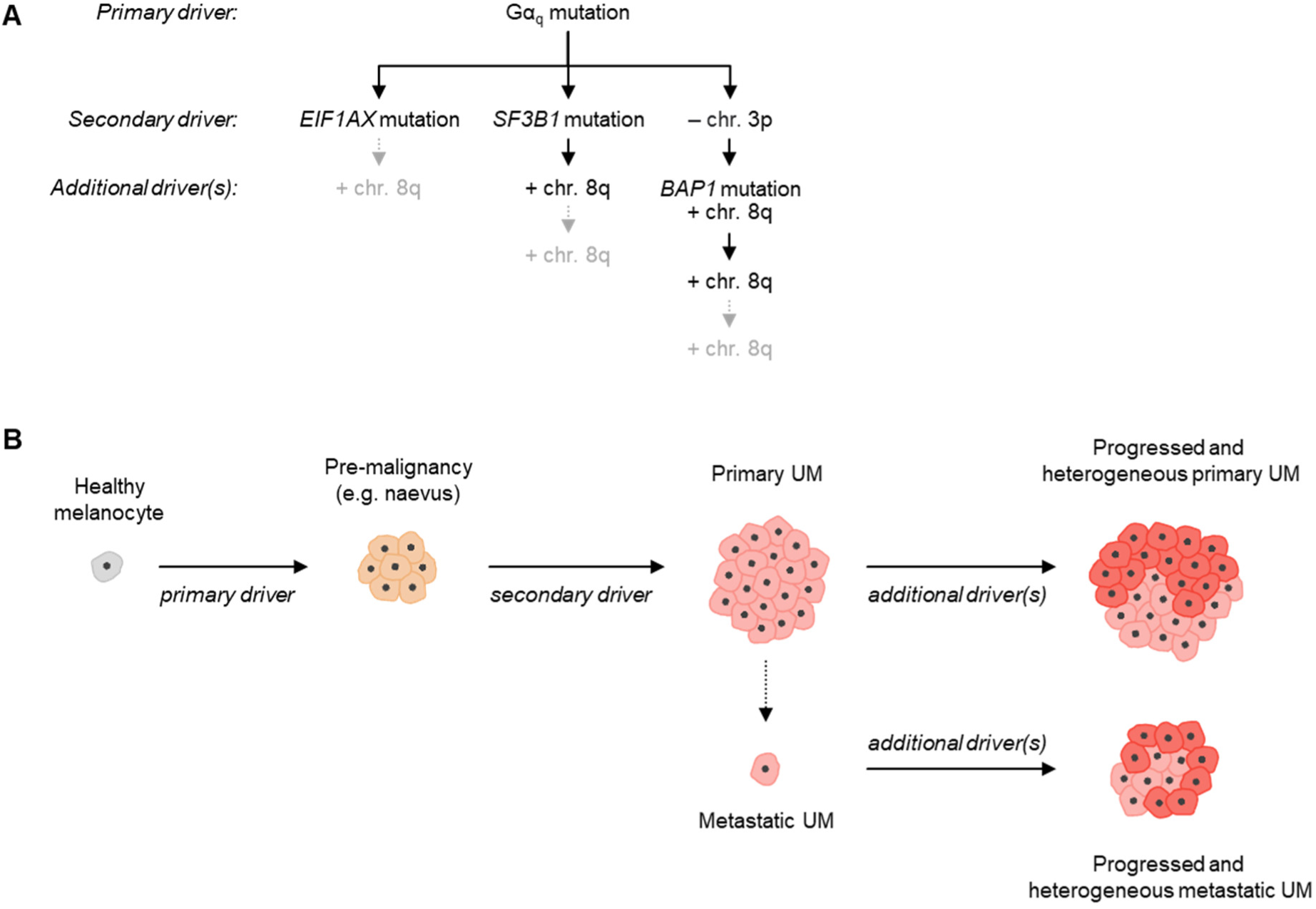
(**A**) Main evolutionary subtypes of UMs with specified primary, secondary and additional genetic alterations driving this evolution. (**B**) Proposed model of gradual tumour evolution ultimately leading to heterogeneous primary and metastatic UMs.

In contrast, a complete and fully clonal inactivation of BAP1 was not found in all tumours with monosomy 3p. This confirms occasional observations in earlier studies [12, 15, 16], and suggests that a somatic mutation in this gene should also be seen as a later evolutionary event. However, as complex *BAP1* alterations can be found throughout the entire gene (**Figure 3A**), not all mutations were quantitatively measured using digital PCR and their clonality could only be analysed in a small subset of the tumours.

We also found that gains and amplifications of chromosome 8q were not restricted to one of the three UM subtypes and frequently showed subclonality. This strongly indicates that these alterations could be seen as a ‘tertiary driver’. Intriguingly, the role and evolutionary timing of chromosome 8q increases has remained largely unclear and controversial until now [15–17, 25]. Using purity-corrected digital PCR-based quantifications, we generated copy number measurements of much higher precision than previous studies. Although some tumours demonstrated an apparently clonal gain or amplification, at least 31% of our cohort presented with a mixture of cells with different 8q copy numbers, making it the most common heterogeneous genetic alteration in primary UM. We confirmed the presence of this heterogeneity in a number of cases using single-cell karyotyping and by analysing a selection of tumour-matched metastatic lesions. These findings suggest a much more complex role for chromosome 8q in UM biology than acknowledged previously.

The progressive ramp-up of 8q copy numbers has been reported as one of the most significant genetic changes observed in the evolution from primary UM to metastasis [16]. Our results, first of all, suggest that the acquisition of additional 8q copies is an ongoing process during progression of the primary tumour: after the primary and secondary drivers have clonally expanded, subclones with increasing 8q copy numbers may appear. Of note, this increase is usually allele-specific, as it typically solely involves the (consecutive) gain of the maternal or paternal allele. Though, it is not entirely clear how this 8q increase in the primary tumour relates to the metastatic progression of UM. Higher 8q copy numbers have been repetitively associated with a higher frequency of and a shorter time to the development of metastatic disease [12, 24, 51–53]. These associations hold true in our current cohort of tumours and may indicate that melanoma cells with higher 8q levels metastasise more easily. On the other hand, UMs are typically large once detected, suggesting a relatively long time of tumour evolution before the primary lesion is treated (and analysed). The increasing 8q copy numbers might then be seen as a ‘molecular clock’ of primary tumour progression, while not necessarily influencing the metastatic potential of the individual cancer cells. In line with this hypothesis, all five metastatic lesions we analysed were supposedly disseminated from a primary tumour clone with disomy 8q. Though, 8q gains or amplifications were present in three of the primary tumours and four of the metastatic lesions, but their allele-specificity and subclonality evidenced that these copy number increases were separate genetic events, occurring independently in the late evolution of both primary and metastatic UM (**Figure 7B**).

Taken together, but in contrast to earlier studies [15, 17], our findings point toward a model of gradual evolution (**Figure 7**) in which a primary driver (i.e. a Gα_q_ signalling mutation) initiates pre-malignant transformation, but a secondary driver (i.e. a mutation in *EIF1AX*, in *SF3B1*, or the loss of chromosome 3p) is required for complete malignant UM development. Subsequently, recurrent tertiary drivers (i.e. most notably chromosome 8q copy number increases) may contribute to full aggressiveness of the cancer, but are not necessary for metastatic dissemination and frequently show heterogeneity within and between tumour lesions in a UM patient.

From a clinical perspective, the presence of any heterogeneity would necessitate a comprehensive and detailed analysis when relying on the detection of genetic alterations in determining the prognosis or follow-up of patients. This specifically applies to the exact (and thus purity-corrected) measurement of the 8q copy number, as this number is a vital parameter in an established classification system of primary UMs [51, 53, 54]. We propose that the digital PCR assays introduced in this study may be an accessible solution to facilitate this analysis. Additionally, our mutation and copy number assays have potential application in sensitively measuring genetic UM patient material of lower quantity or quality, such as small solid or liquid biopsies. This is illustrated by the successful analysis of FFPE-derived metastases in this study. As another example, some of the mutation assays have already been used in blood-based screening for circulating tumour DNA [55], and the clinical value of similar applications is increasingly recognised [56].

Critically, our current study is limited by the sole analysis of primary tumours obtained from enucleated eyes, making our cohort being biased toward larger and possibly more evolved (and more clonal) tumours. We hypothesise that the earliest stages of tumour evolution may be studied by comparable in-depth analyses of smaller UMs. In addition, it would be interesting to investigate more pairs of primary and metastatic UMs to better define the later stages of UM evolution. Our evaluations may be further extended to the analysis of other recurrent or sample-specific genetic alterations present in UM.

In conclusion, using novel digital PCR-based approaches, we identified systematic differences in the evolutionary timing of the key genetic events in UM. The observed intratumour heterogeneity suggests a more complex model of gradual tumour evolution and argues for a comprehensive genetic analysis in clinical practice, which may be facilitated by the sensitive digital PCR assays developed in this study.

## Data Availability

All data produced in the present work are contained in the manuscript.

https://github.com/rjnell/um-heterogeneity

## Acknowledgements

This study was supported by the European Union’s Horizon 2020 research and innovation program under grant agreement number 667787 (UM Cure 2020, R.J. Nell and N.V. Menger). We thank D. Cats, H. Mei, L. van Vught and M.C. Gelmi for their help with the sequencing data and clinical information from the patients in this study.

## Notes

### Competing Interest Statement

The authors have declared no competing interest.

### Funding Statement

This study was funded by the European Union's Horizon 2020 research and innovation program under grant agreement number 667787 (UM Cure 2020, R.J. Nell and N.V. Menger).

### Author Declarations

This study was approved by the Leiden University Medical Center Biobank Committee and Medisch Ethische Toetsingscommissie under no. B14.003/DH/sh and B20.026.

## References

1. Coupland, S.E., et al., Molecular pathology of uveal melanoma. Eye (Lond), 2013. 27(2): p. 230–42.

2. Jager, M.J., et al., Uveal melanoma. Nat Rev Dis Primers, 2020. 6(1): p. 24.

3. Furney, S.J., et al., SF3B1 mutations are associated with alternative splicing in uveal melanoma. Cancer Discov, 2013. 3(10): p. 1122–1129.

4. Royer-Bertrand, B., et al., Comprehensive Genetic Landscape of Uveal Melanoma by Whole-Genome Sequencing. Am J Hum Genet, 2016. 99(5): p. 1190–1198.

5. Van Raamsdonk, C.D., et al., Frequent somatic mutations of GNAQ in uveal melanoma and blue naevi. Nature, 2009. 457(7229): p. 599–602.

6. Van Raamsdonk, C.D., et al., Mutations in GNA11 in uveal melanoma. N Engl J Med, 2010. 363(23): p. 2191–9.

7. Johansson, P., et al., Deep sequencing of uveal melanoma identifies a recurrent mutation in PLCB4. Oncotarget, 2016. 7(4): p. 4624–31.

8. Moore, A.R., et al., Recurrent activating mutations of G-protein-coupled receptor CYSLTR2 in uveal melanoma. Nat Genet, 2016. 48(6): p. 675–80.

9. Harbour, J.W., et al., Frequent mutation of BAP1 in metastasizing uveal melanomas. Science, 2010. 330(6009): p. 1410–3.

10. Harbour, J.W., et al., Recurrent mutations at codon 625 of the splicing factor SF3B1 in uveal melanoma. Nat Genet, 2013. 45(2): p. 133–5.

11. Martin, M., et al., Exome sequencing identifies recurrent somatic mutations in EIF1AX and SF3B1 in uveal melanoma with disomy 3. Nat Genet, 2013. 45(8): p. 933–6.

12. Robertson, A.G., et al., Integrative Analysis Identifies Four Molecular and Clinical Subsets in Uveal Melanoma. Cancer Cell, 2017. 32(2): p. 204–220 e15.

13. Yavuzyigitoglu, S., et al., Uveal Melanomas with SF3B1 Mutations: A Distinct Subclass Associated with Late-Onset Metastases. Ophthalmology, 2016. 123(5): p. 1118–28.

14. Onken, M.D., et al., Gene expression profiling in uveal melanoma reveals two molecular classes and predicts metastatic death. Cancer Res, 2004. 64(20): p. 7205–9.

15. Field, M.G., et al., Punctuated evolution of canonical genomic aberrations in uveal melanoma. Nat Commun, 2018. 9(1): p. 116.

16. Shain, A.H., et al., The genetic evolution of metastatic uveal melanoma. Nat Genet, 2019. 51(7): p. 1123–1130.

17. Rodrigues, M., et al., Evolutionary Routes in Metastatic Uveal Melanomas Depend on MBD4 Alterations. Clin Cancer Res, 2019. 25(18): p. 5513–5524.

18. Caravagna, G., et al., Subclonal reconstruction of tumors by using machine learning and population genetics. Nat Genet, 2020. 52(9): p. 898–907.

19. Durante, M.A., et al., Single-cell analysis reveals new evolutionary complexity in uveal melanoma. Nat Commun, 2020. 11(1): p. 496.

20. Christodoulou, E., et al., Loss of Wild-Type CDKN2A Is an Early Event in the Development of Melanoma in FAMMM Syndrome. J Invest Dermatol, 2020. 140(11): p. 2298–2301 e3.

21. Nell, R.J., et al., Involvement of mutant and wild-type CYSLTR2 in the development and progression of uveal nevi and melanoma. BMC Cancer, 2021. 21(1): p. 164.

22. van Eijk, R., et al., Assessment of a fully automated high-throughput DNA extraction method from formalin-fixed, paraffin-embedded tissue for KRAS, and BRAF somatic mutation analysis. Exp Mol Pathol, 2013. 94(1): p. 121–5.

23. Nell, R.J., et al., Identification of clinically-relevant genetic alterations in uveal melanoma using RNA sequencing. 2023.

24. Versluis, M., et al., Digital PCR validates 8q dosage as prognostic tool in uveal melanoma. PLoS One, 2015. 10(3): p. e0116371.

25. de Lange, M.J., et al., Heterogeneity revealed by integrated genomic analysis uncovers a molecular switch in malignant uveal melanoma. Oncotarget, 2015. 6(35): p. 37824–35.

26. Zoutman, W.H., R.J. Nell, and P.A. van der Velden, Usage of Droplet Digital PCR (ddPCR) Assays for T Cell Quantification in Cancer. Methods Mol Biol, 2019. 1884: p. 1–14.

27. Nell, R.J., et al., Allele-specific digital PCR enhances precision and sensitivity in the detection and quantification of copy number alterations in heterogeneous DNA samples: an in silico and in vitro validation study. medRxiv, 2023: p. 2023.10.31.23297362.

28. Dube, S., J. Qin, and R. Ramakrishnan, Mathematical analysis of copy number variation in a DNA sample using digital PCR on a nanofluidic device. PLoS One, 2008. 3(8): p. e2876.

29. Smit, K.N., et al., Combined mutation and copy-number variation detection by targeted next-generation sequencing in uveal melanoma. Mod Pathol, 2018. 31(5): p. 763–771.

30. van Essen, T.H., et al., Prognostic parameters in uveal melanoma and their association with BAP1 expression. Br J Ophthalmol, 2014. 98(12): p. 1738–43.

31. Johnson, C.P., et al., Systematic genomic and translational efficiency studies of uveal melanoma. PLoS One, 2017. 12(6): p. e0178189.

32. Johansson, P.A., et al., Whole genome landscapes of uveal melanoma show an ultraviolet radiation signature in iris tumours. Nat Commun, 2020. 11(1): p. 2408.

33. Bidshahri, R., et al., Quantitative Detection and Resolution of BRAF V600 Status in Colorectal Cancer Using Droplet Digital PCR and a Novel Wild-Type Negative Assay. J Mol Diagn, 2016. 18(2): p. 190–204.

34. Alsafadi, S., et al., Cancer-associated SF3B1 mutations affect alternative splicing by promoting alternative branchpoint usage. Nat Commun, 2016. 7: p. 10615.

35. Karlsson, J., et al., Molecular profiling of driver events in metastatic uveal melanoma. Nat Commun, 2020. 11(1): p. 1894.

36. Durinck, S., et al., Temporal dissection of tumorigenesis in primary cancers. Cancer Discov, 2011. 1(2): p. 137–43.

37. Nik-Zainal, S., et al., The life history of 21 breast cancers. Cell, 2012. 149(5): p. 994–1007.

38. Mitchell, T.J., et al., Timing the Landmark Events in the Evolution of Clear Cell Renal Cell Cancer: TRACERx Renal. Cell, 2018. 173(3): p. 611–623 e17.

39. Gerstung, M., et al., The evolutionary history of 2,658 cancers. Nature, 2020. 578(7793): p. 122–128.

40. Martincorena, I. and P.J. Campbell, Somatic mutation in cancer and normal cells. Science, 2015. 349(6255): p. 1483–9.

41. Jolly, C. and P. Van Loo, Timing somatic events in the evolution of cancer. Genome Biol, 2018. 19(1): p. 95.

42. Gerlinger, M., et al., Intratumor heterogeneity and branched evolution revealed by multiregion sequencing. N Engl J Med, 2012. 366(10): p. 883–892.

43. Shain, A.H., et al., The Genetic Evolution of Melanoma from Precursor Lesions. N Engl J Med, 2015. 373(20): p. 1926–36.

44. Jamal-Hanjani, M., et al., Tracking the Evolution of Non-Small-Cell Lung Cancer. N Engl J Med, 2017. 376(22): p. 2109–2121.

45. Shain, A.H., et al., Genomic and Transcriptomic Analysis Reveals Incremental Disruption of Key Signaling Pathways during Melanoma Evolution. Cancer Cell, 2018. 34(1): p. 45–55 e4.

46. Vader, M.J.C., et al., GNAQ and GNA11 mutations and downstream YAP activation in choroidal nevi. Br J Cancer, 2017. 117(6): p. 884–887.

47. Durante, M.A., et al., Genomic evolution of uveal melanoma arising in ocular melanocytosis. Cold Spring Harb Mol Case Stud, 2019. 5(4).

48. Silva-Rodriguez, P., et al., GNAQ and GNA11 Genes: A Comprehensive Review on Oncogenesis, Prognosis and Therapeutic Opportunities in Uveal Melanoma. Cancers (Basel), 2022. 14(13).

49. Maat, W., et al., The heterogeneous distribution of monosomy 3 in uveal melanomas: implications for prognostication based on fine-needle aspiration biopsies. Arch Pathol Lab Med, 2007. 131(1): p. 91–6.

50. Mensink, H.W., et al., Chromosome 3 intratumor heterogeneity in uveal melanoma. Invest Ophthalmol Vis Sci, 2009. 50(2): p. 500–4.

51. Vichitvejpaisal, P., et al., Genetic Analysis of Uveal Melanoma in 658 Patients Using the Cancer Genome Atlas Classification of Uveal Melanoma as A, B, C, and D. Ophthalmology, 2019. 126(10): p. 1445–1453.

52. Thornton, S., et al., Targeted Next-Generation Sequencing of 117 Routine Clinical Samples Provides Further Insights into the Molecular Landscape of Uveal Melanoma. Cancers (Basel), 2020. 12(4).

53. Shields, C.L., et al., Ten-year outcomes of uveal melanoma based on The Cancer Genome Atlas (TCGA) classification in 1001 cases. Indian J Ophthalmol, 2021. 69(7): p. 1839–1845.

54. Jager, M.J., N.J. Brouwer, and B. Esmaeli, The Cancer Genome Atlas Project: An Integrated Molecular View of Uveal Melanoma. Ophthalmology, 2018. 125(8): p. 1139–1142.

55. Beasley, A., et al., Clinical Application of Circulating Tumor Cells and Circulating Tumor DNA in Uveal Melanoma. JCO Precis Oncol, 2018. 2.

56. Carvajal, R.D., et al., Clinical and molecular response to tebentafusp in previously treated patients with metastatic uveal melanoma: a phase 2 trial. Nat Med, 2022. 28(11): p. 2364–2373.

