## Supplementary Information for "Digital PCR-based deep quantitative profiling delineates heterogeneity and evolution of uveal melanoma"

### Supplementary Table 1

Overview of clinical and molecular characteristics of all cases analysed in this study.

### Supplementary Table 2

Context sequence, annealing temperature and available supplier information for digital PCR assays used in this study.

**Target assays:**

| Assay | *PPARG* target chromosome 3p (FAM-labelled) |
| --- | --- |
| Location (hg38) | chr3:12381327-12381449 |
| Amplicon length | 69 nucleotides |
| Annealing temperature | 60 °C |
| Supplier and assay ID | Bio-Rad (dHsaCP100462) |

| Assay | *PTK2* target chromosome 8q (FAM-labelled) |
| --- | --- |
| Location (hg38) | chr8:140659331-140659453 |
| Amplicon length | 70 nucleotides |
| Annealing temperature | 60 °C |
| Supplier and assay ID | Bio-Rad (dHsaCP1000155) |

**Reference assays:**

| Assay | *TERT* reference chromosome 5p (HEX-labelled) |
| --- | --- |
| Location (hg38) | chr5:1258577-1258699 |
| Amplicon length | 100 nucleotides |
| Annealing temperature | 60 °C |
| Supplier and assay ID | Bio-Rad (dHsaCP1000100) |

| Assay | *VOPP1* reference chromosome 7p (FAM-labelled) |
| --- | --- |
| Location (hg38) | chr7:55497633-55497755 |
| Amplicon length | 64 nucleotides |
| Annealing temperature | 60 °C |
| Supplier and assay ID | Bio-Rad (dHsaCP2506292) |

| Assay | *BRAF* reference chromosome 7q (HEX-labelled) |
| --- | --- |
| Location (hg38) | chr7:140800362-140800484 |
| Amplicon length | 66 nucleotides |
| Annealing temperature | 60 °C |
| Supplier and assay ID | Bio-Rad (dHsaCP2500366) |

| Assay | *TTC5* reference chromosome 14q (HEX-labelled) |
| --- | --- |
| Location (hg38) | chr14:20289639-20289761 |
| Amplicon length | 59 nucleotides |
| Annealing temperature | 60 °C |
| Supplier and assay ID | Bio-Rad (dHsaCP2506733) |

**SNP assays:**

| Assay | SNP rs6617 (G/C) chromosome 3p (FAM/HEX-labelled) |
| --- | --- |
| Location (hg38) | chr3:52706096-52706245 |
| Amplicon length | 63 nucleotides |
| Annealing temperature | 55 °C |
| Supplier | Sigma-Aldrich |

| Assay (supplier) | SNP rs6976 (T/C) chromosome 3p (FAM/HEX-labelled) |
| --- | --- |
| Location (hg38) | chr3:52694726-52694848 |
| Amplicon length | 77 nucleotides |
| Annealing temperature | 55 °C |
| Supplier | Sigma-Aldrich |

| Assay (supplier) | SNP rs9586 (T/C) chromosome 3p (FAM/HEX-labelled) |
| --- | --- |
| Location (hg38) | chr3:49176159-49176292 |
| Amplicon length | 84 nucleotides |
| Annealing temperature | 55 °C |
| Supplier | Sigma-Aldrich |

| Assay (supplier) | SNP rs1989839 (G/A) chromosome 3p (FAM/HEX-labelled) |
| --- | --- |
| Location (hg38) | chr3:50341454-50341576 |
| Amplicon length | 69 nucleotides |
| Annealing temperature | 55 °C |
| Supplier | Bio-Rad (dHsaMDS621043799) |

| Assay | SNP rs1062633 (C/T) chromosome 3p (FAM/HEX-labelled) |
| --- | --- |
| Location (hg38) | chr3:49887436-49887620 |
| Amplicon length | 130 nucleotides |
| Annealing temperature | 55 °C |
| Supplier | Sigma-Aldrich |

| Assay | SNP rs2236947 (C/A) chromosome 3p (FAM/HEX-labelled) |
| --- | --- |
| Location (hg38) | chr3:50333940-50334062 |
| Amplicon length | 69 nucleotides |
| Annealing temperature | 55 °C |
| Supplier and assay ID | Bio-Rad (dHsaMDS958483376) |

| Assay | SNP rs7018178 (C/T) chromosome 8q (FAM/HEX-labelled) |
| --- | --- |
| Location (hg38) | chr8:141229028-141229207 |
| Amplicon length | 122 nucleotides |
| Annealing temperature | 55 °C |
| Supplier | Sigma-Aldrich |

| Assay | SNP rs7843014 (A/C) chromosome 8q (FAM/HEX-labelled) |
| --- | --- |
| Location (hg38) | chr8:140780306-140780445 |
| Amplicon length | 112 nucleotides |
| Annealing temperature | 55 °C |
| Supplier | Sigma-Aldrich |

**Targeted mutation assays:**

| Assay | *GNAQ* p.G48L mutation/wildtype (FAM/HEX-labelled) |
| --- | --- |
| Location (hg38) | chr9:77922272-77922409 |
| Amplicon length | 102 nucleotides |
| Annealing temperature | 55 °C |
| Supplier | Sigma-Aldrich |

| Assay | *GNAQ* p.Q209P/L mutation/wildtype (FAM/HEX-labelled) |
| --- | --- |
| Location (hg38) | chr9:77794511-77794633 |
| Amplicon length | 65 nucleotides |
| Annealing temperature | 55 °C |
| Supplier and assay IDs | Bio-Rad (dHsaCP2000051/52 and dHsaCP250676794) |

| Assay | *GNA11* p.Q209P/L mutation/wildtype (FAM/HEX-labelled) |
| --- | --- |
| Location (hg38) | chr19:3118883-3119005 |
| Amplicon length | 62 nucleotides |
| Annealing temperature | 55 °C |
| Supplier and assay ID | Bio-Rad (dHsaCP2000049/50) |

| Assay | *CYSLTR2* p.L129Q mutation/wildtype (FAM/HEX-labelled) |
| --- | --- |
| Location (hg38) | chr13:48707095-48707274 |
| Amplicon length | 101 nucleotides |
| Annealing temperature | 55 °C |
| Supplier | Sigma-Aldrich |

| Assay | *GNA11* p.R183C/H/Y mutation/wildtype (FAM/HEX-labelled) |
| --- | --- |
| Location (hg38) | chr19:3114941-3115074 |
| Amplicon length | 88 nucleotides |
| Annealing temperature | 55 °C |
| Supplier | Sigma-Aldrich |

| Assay | *PLCB4* p.D630F/N mutation/wildtype (FAM/HEX-labelled) |
| --- | --- |
| Location (hg38) | chr20:9409034-9409168 |
| Amplicon length | 88 nucleotides |
| Annealing temperature | 55 °C |
| Supplier | Sigma-Aldrich |

| Assay | *SF3B1* p.R625C/H mutation/wildtype (FAM/HEX-labelled) |
| --- | --- |
| Location (hg38) | chr2:197402692-197402849 |
| Amplicon length | 112 nucleotides |
| Annealing temperature | 55 °C |
| Supplier | Sigma-Aldrich |

| Assay | *BAP1* exon 4 mutation/wildtype (FAM/HEX-labelled) |
| --- | --- |
| Location (hg38) | chr3:52408443-52408579 |
| Amplicon length | 85 nucleotides |
| Annealing temperature | 55 °C |
| Supplier | Sigma-Aldrich |

| Assay | *BAP1* intron 5 mutation/wildtype (FAM/HEX-labelled) |
| --- | --- |
| Location (hg38) | chr3:52407404-52407547 |
| Amplicon length | 86 nucleotides |
| Annealing temperature | 55 °C |
| Supplier | Sigma-Aldrich |

| Assay | *BAP1* exon 7 mutation/wildtype (FAM/HEX-labelled) |
| --- | --- |
| Location (hg38) | chr3:52407121-52407278 |
| Amplicon length | 102 nucleotides |
| Annealing temperature | 55 °C |
| Supplier | Sigma-Aldrich |

| Assay | *BAP1* exon 7 (alt.) mutation/wildtype (FAM/HEX-labelled) |
| --- | --- |
| Location (hg38) | chr3:52407121-52407317 |
| Amplicon length | 147 nucleotides |
| Annealing temperature | 55 °C |
| Supplier | Sigma-Aldrich |

| Assay | *BAP1* intron 8 mutation/wildtype (FAM/HEX-labelled) |
| --- | --- |
| Location (hg38) | chr3:52406846-52406992 |
| Amplicon length | 91 nucleotides |
| Annealing temperature | 55 °C |
| Supplier | Sigma-Aldrich |

| Assay | *BAP1* exon 9 mutation/wildtype (FAM/HEX-labelled) |
| --- | --- |
| Location (hg38) | chr3:52406270-52406409 |
| Amplicon length | 98 nucleotides |
| Annealing temperature | 55 °C |
| Supplier | Sigma-Aldrich |

| Assay | *BAP1* exon 11 mutation/wildtype (FAM/HEX-labelled) |
| --- | --- |
| Location (hg38) | chr3:52405141-52405308 |
| Amplicon length | 128 nucleotides |
| Annealing temperature | 55 °C |
| Supplier | Sigma-Aldrich |

| Assay | *BAP1* exon 15 mutation/wildtype (FAM/HEX-labelled) |
| --- | --- |
| Location (hg38) | chr3:52402795-52402926 |
| Amplicon length | 86 nucleotides |
| Annealing temperature | 55 °C |
| Supplier | Sigma-Aldrich |

**Drop-off mutation assays:**

| Assay | *EIF1AX* exon 1 (FAM/HEX-labelled) |
| --- | --- |
| Location (hg38) | chrX:20141585-20141706 |
| Amplicon length | 82 nucleotides |
| Annealing temperature | 55 °C |
| Supplier | Sigma-Aldrich |

| Assay | *EIF1AX* exon 2 (FAM/HEX-labelled) |
| --- | --- |
| Location (hg38) | chrX:20138523-20138699 |
| Amplicon length | 123 nucleotides |
| Annealing temperature | 58 °C |
| Supplier | Sigma-Aldrich |

| Assay | *SF3B1* exon 14 (FAM/HEX-labelled) |
| --- | --- |
| Location (hg38) | chr2:197402577-197402849 |
| Amplicon length | 227 nucleotides |
| Annealing temperature | 55 °C |
| Supplier | Sigma-Aldrich |

| Assay | *BAP1* exon 5 (FAM/HEX-labelled) |
| --- | --- |
| Location (hg38) | chr3:52407994-52408129 |
| Amplicon length | 95 nucleotides |
| Annealing temperature | 55 °C |
| Supplier | Sigma-Aldrich |

### Supplementary Figure 1

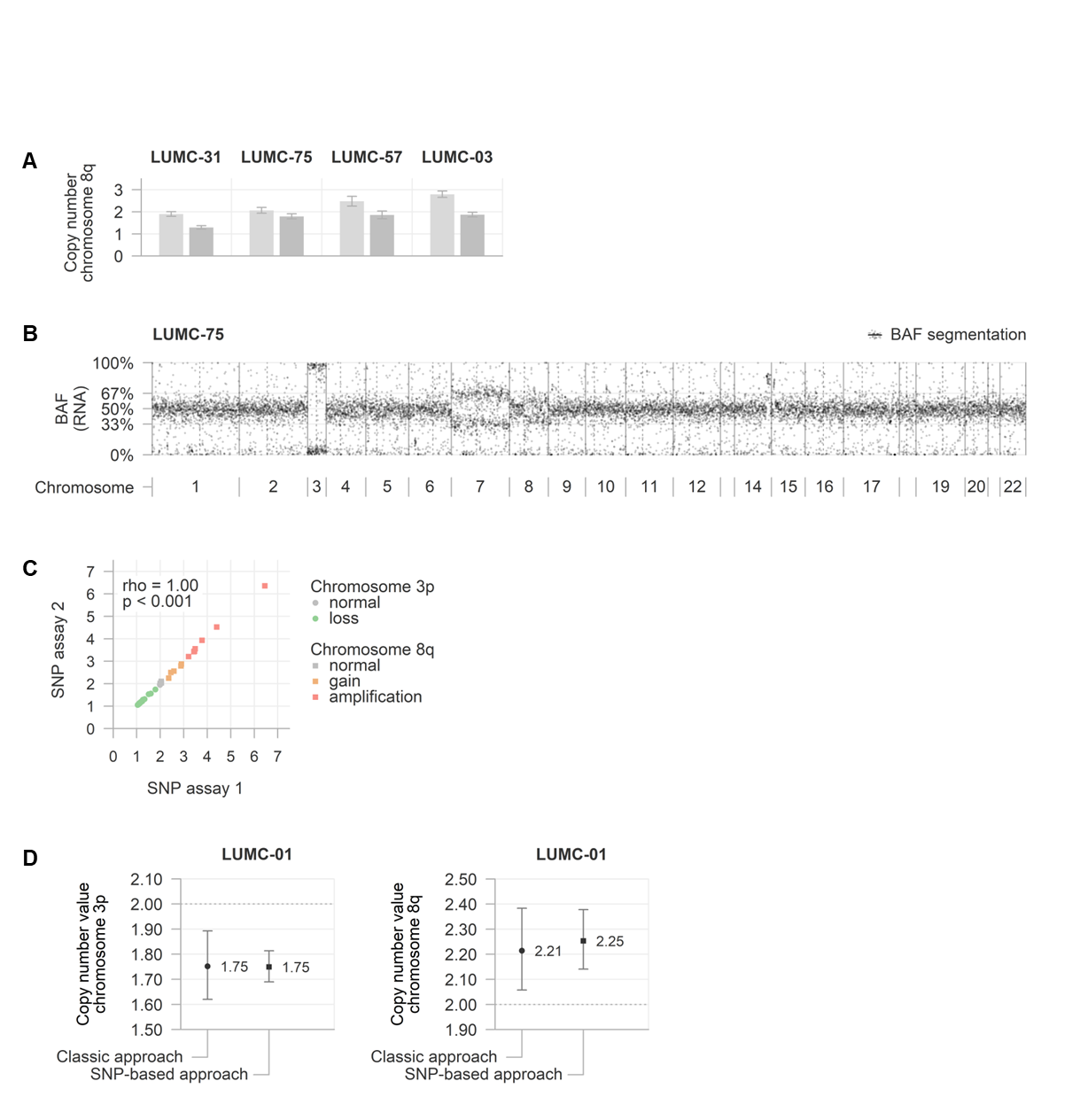

(**A**) Allele-specific copy number values of tumours with biallelic chromosome 8q copy number increases, extended from **Figure 4B**.

(**B**) RNA-inferred allelic imbalances showing imbalanced expression of a limited number of chromosomes, including 3, 7 and 8q.

(**C**) Comparison between copy number values determined with multiple SNP assays.

(**D**) Example of enhanced precision of the SNP-based approach in comparison to the classic approach in the measurement of chromosome 3p and 8q copy number values.

### Supplementary Figure 2

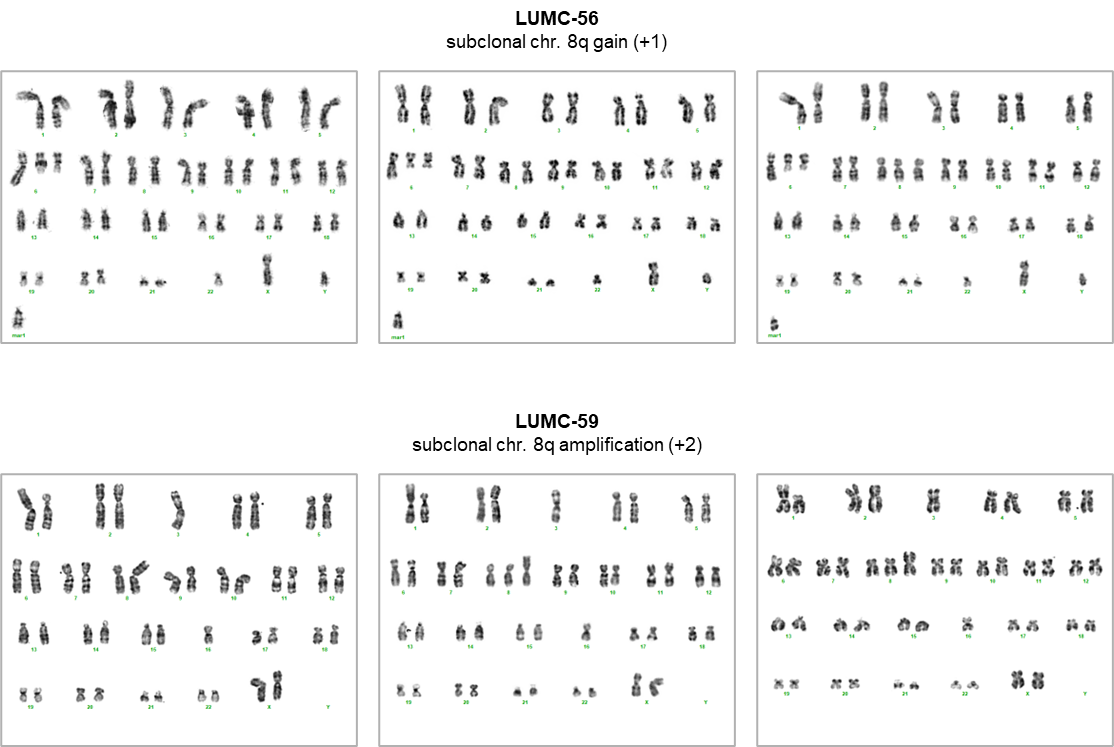

Complete karyograms of LUMC-56 and -59 (as presented in **Figure 5B**), demonstrating heterogeneity with regard to the copy number of chromosome 8q.
